# Quantitative Method for Comparative Assessment of Particle Filtration Efficiency of Fabric Masks as Alternatives to Standard Surgical Masks for PPE

**DOI:** 10.1101/2020.04.17.20069567

**Authors:** Amy V. Mueller, Matthew J. Eden, Jessica M. Oakes, Chiara Bellini, Loretta A. Fernandez

## Abstract

In response to the COVID-19 pandemic, cloth masks are being used to control the spread of virus, but the efficacy of these loose-fitting masks is not well known. Here, tools and methods typically used to assess tight-fitting respirators were modified to quantify the efficacy of community- and commercially-produced fabric masks as PPE. Two particle counters concurrently sample ambient air and air inside the masks; mask performance is evaluated by mean particle removal efficiency and statistical variability when worn as designed and with a nylon overlayer, to independently assess fit and material. Worn as designed both commercial surgical masks and cloth masks had widely varying effectiveness (53-75% and 28-90% filtration efficiency, respectively). Most surgical-style masks improved with the nylon overlayer, indicating poor fit. This rapid testing method uses widely available hardware, requires only a few calculations from collected data, and provides both a holistic and aspect-wise evaluation of mask performance.

## 1. Introduction

In response to the critical shortage of medical masks resulting from the COVID-19 pandemic, large portions of the population are mobilizing to produce cloth masks using locally-sourced fabrics. While the general population is being advised to wear masks to protect others from virus that may be spread from the wearer, the efficacy of these masks as a means of protecting the wearer from airborne particles carrying virus is also a concern, particularly as medical masks grow scarce. This issue may become more critical if it becomes necessary for medical care workers to use similar alternative personal protective equipment, but is already important for individuals who may be caring for a household member who is ill or who may be in a high-risk category for complications.^2^

The effectiveness of masks to protect wearers from airborne particles is known to be a function of both materials and fit. Standard methods to test the performance of respirators and masks designed to form a seal against the face, such as N95 respirators, assume that appropriate high-filtration materials have been used in the construction of the masks and therefore employ instruments that test the fit by comparing the concentration of particles in air inside and outside of the mask while the subject moves his/her head through a series of positions.^3^ Several instruments have been specifically designed to perform these tests (e.g., the TSI PortaCount), simplifying the testing process for users by reporting a single metric of “fit” (i.e., Fit Factor = ratio of time-averaged particle concentration outside and inside mask). In contrast, standard methods for surgical masks focus exclusively on testing the materials and do not provide for a measurement of the mask as constructed or as worn.^4-7^

Anticipating the need to produce face coverings from readily-available materials, several studies have used these standard methods for materials testing to compare the filtration efficiency of materials such as cotton t-shirts, sweatshirts, handkerchiefs, and towels with the filtration efficiency of materials used to manufacture facepiece respirators (N95 masks) and surgical masks.^8-10^ Further, tools developed for N95-type masks have been applied directly to evaluate particle filtration for loose-fitting, surgical type masks.^11^ Generally, these studies have found that no commonly-available materials produce filtration efficiency close to respirators such as N95s, with cotton cloth facemasks providing about half the protection (i.e., “Fit Factor” decrease by a factor of 2) of standard surgical masks against airborne particles.^11^ Notably, these previous studies are unable to pinpoint the problems with loose-fitting masks (i.e., separate out a poor fit from poor materials used in the construction), though in other work an elastic layer (e.g., nylon stocking) placed over the mask when worn has been found to improve filtration efficiency of loose-fitting masks by minimizing air flow around the cloth layers.^12^

Importantly, it has been shown that head motions and positions do not significantly affect the performance of loose-fitting masks in terms of filtering out nano-sized particles,^11^ suggesting that a simplified mask testing protocol (compared to the multi-step fit test used for respirators) may be sufficient for characterizing particle filtration efficacy of loose-fitting masks. Given the highly varied results and protocol shortcomings noted for prior studies,^13^ development of a rapid and quantitative method for evaluating potential PPE options would be of great value to the general public at this time.

The purpose of this work was to develop a standardized method to quantitatively assess the efficacy of sewn fabric facemasks and standard surgical masks in terms of protecting the wearer from airborne particulates of the size range potentially associated with viral transmission (<300 nm). This leverages instrumentation designed for respirator fit testing, which is widely available nationally at health care centers, fire departments, etc., but provides two key adjustments that improve the data quality for loose-fitting mask testing. First, two instruments are used to simultaneously record high resolution (1 Hz) particle concentration measurements in the room and behind the mask, enabling the method to be used in cases where particle concentration may vary on the timescale of tests, in comparison to standard fit testing which assumes consistent particle concentrations in the room over ~ minutes and therefore sequentially samples the ambient and in-mask air. Data recorded during experiments described below show variability of particle concentrations by up to a factor of 2 over <1 minute, supporting the need for this dual-instrument configuration if used more broadly, especially outside of specialized testing rooms. Second, by conducting separate tests for masks worn loosely (as designed) and for the masks held closeto the face using a layer of nylon stocking (as recommended by Cooper et al.^12^), the method enables independent evaluation of the mask fit and mask materials as they contribute to overall filtration efficiency.

The proposed protocol enables testing of an individual mask design (n=3 masks for statistical analysis) within ~30 minutes for systems with digital data collection, providing a rapid screening tool to test a variety of mask designs produced from readily-available materials. This method can be easily replicated in health care centers, fire stations, and other facilities nationally, to vet specific masks in near-real time. To validate the methodology, this manuscript reports data collected from an initial set of commercial and homemade masks, however results from ongoing tests are being updated regularly at a public web portal as additional prototype masks are evaluated. Given the limited time and current social distancing precautions, all tests were conducted while masks were being worn by the same subject, breathing normally, through the nose, with the mouth closed, while holding the head at a steady position. Data reported by van der Sande et al.^11^ provide confidence that limitation of motions and positions does not significantly limit the conclusions that can be drawn from the resulting data, and results from a single test subject are used here primarily to validate the protocol itself.

## 2. Results and Discussion

The percent removal of particles (of size range characterized below, D<300 nm) for each mask was computed from data collected each second over one minute tests; examples of the high resolution data collected for each test are provided in Figure 1 for a well-fitted N95 mask (N95-1) and a surgical-style cloth mask (CS-1). The breathing pattern of the wearer can be observed as oscillations in the “inside mask” data, and the issue of variability in ambient particle concentrations over a 1-minute test is clearly visible in the top panel.

**Figure 1:**
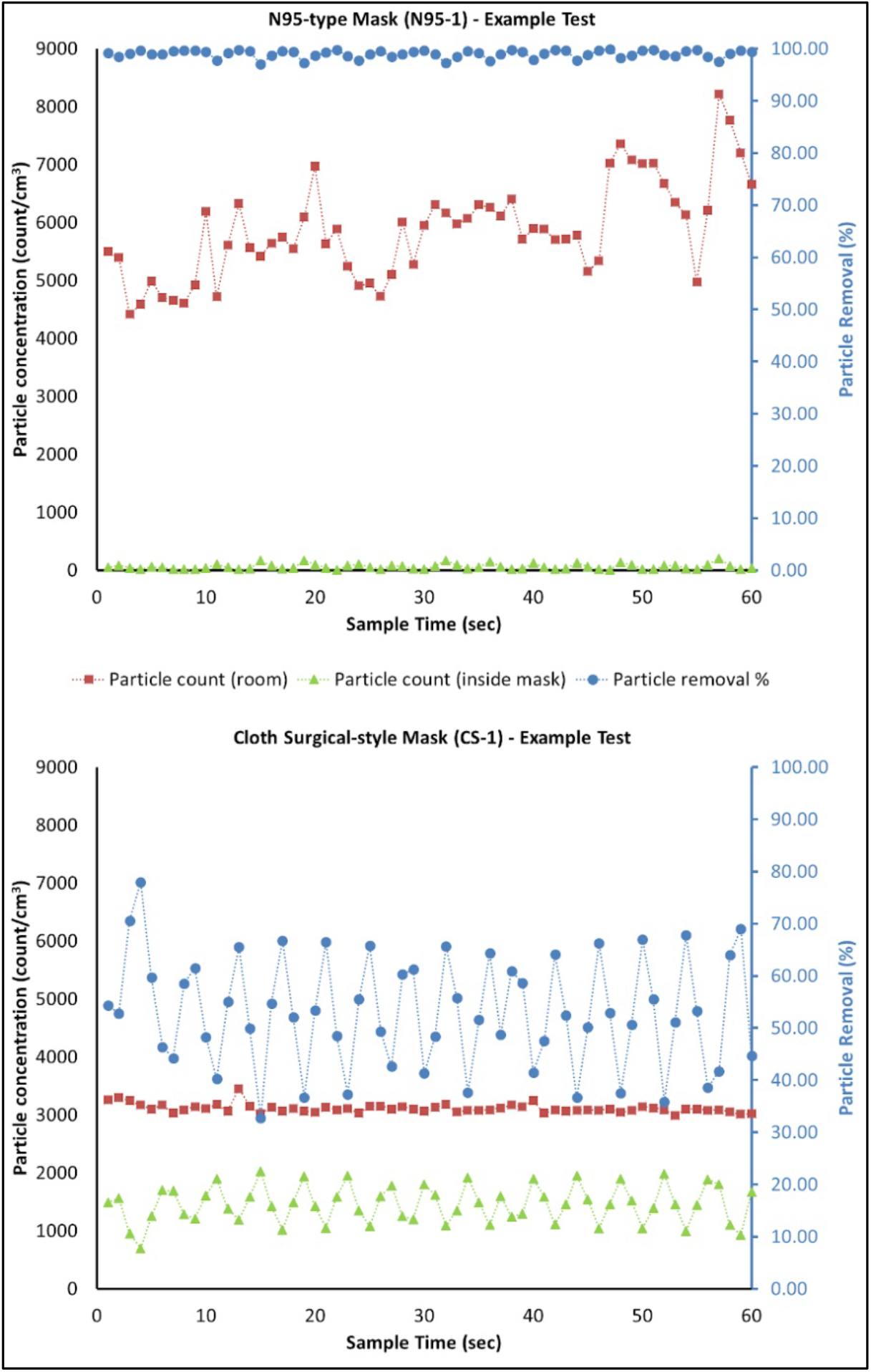
Particle concentrations in the room (red squares) and inside the mask (green triangles) with calculated removal percentage (blue circles) vs. time for a single one-minute test of a well-fitted N95 mask (N95-1, top) and an example cloth surgical-style mask (CS-1, bottom). Time-based variability in filtration efficiency corresponds to the breathing patterns of the mask wearer (inhales vs. exhales). As expected, the N95 mask has high and consistent filtration efficiency (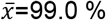, *s_t_*=0.75 % for this single test). The cloth surgical-style mask has both lower filtration efficiency and higher variability (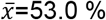, *s_t_*=10.5 % for this single test).

From these data, one can extract both mean removal efficiency and a measure of time-based variation (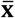 and *s_t_*, as defined below), which each provide information on mask performance. It is observed that 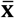 and *s_t_* are inversely correlated (Figure 2), wherein an improved fit generally leads to both higher mean particle removal efficiency and lower time-based standard deviation (consistency in particle removal), independent of the materials being used.

**Figure 2:**
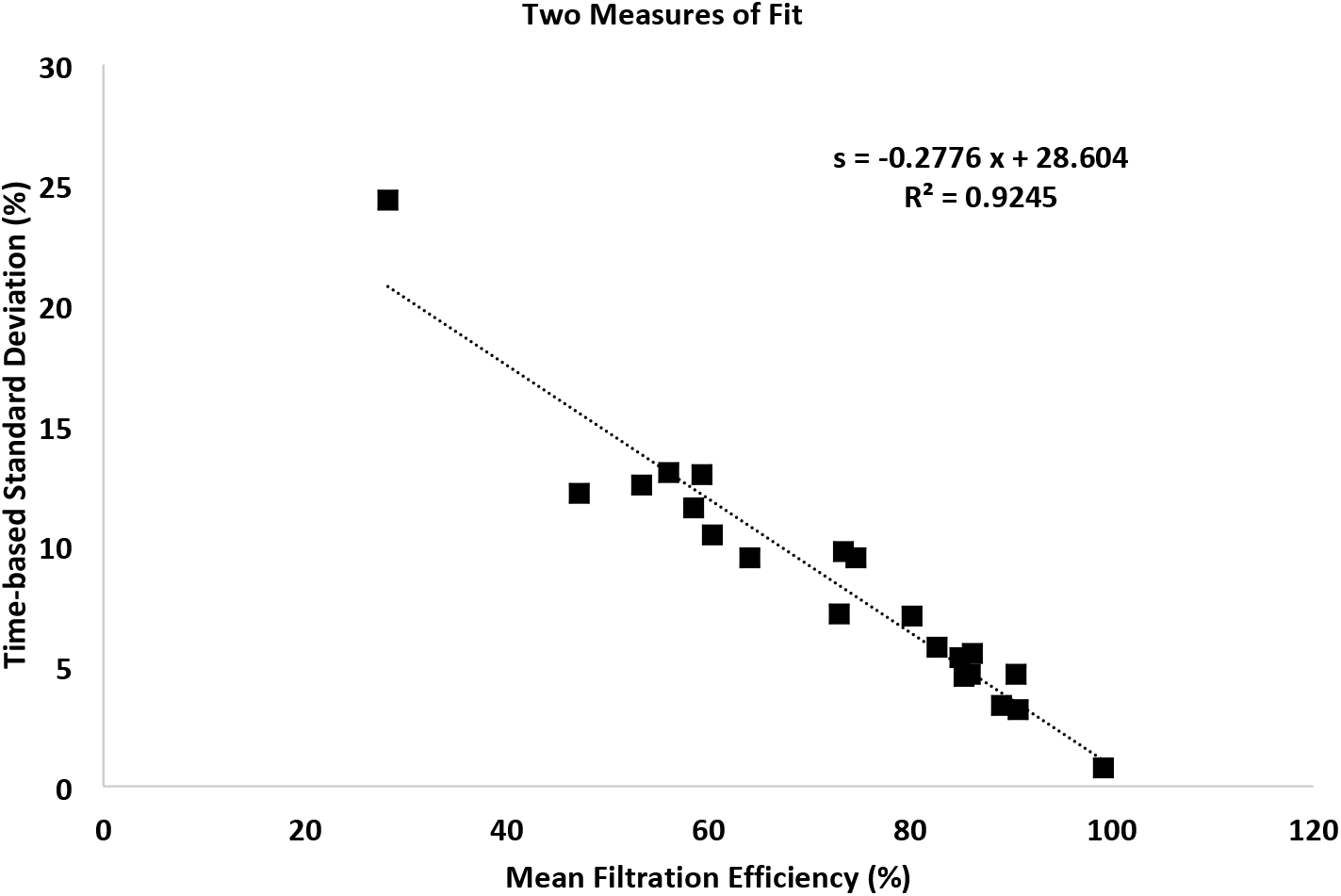
Improving mask performance through better fit and filtration materials leads both to increased mean particle removal efficiency and decreased variation in filtration overtime; data shown for masks worn as designed.

Data collected with the subject wearing the nylon overlayer alone had 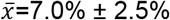 (standard deviation calculated from n=3 replicates) with *s_t_*=18%; it is concluded therefore that the overlayer itself does not provide significant filtration capacity and in the following discussion it is considered primarily to improve the snugness of fit of the underlying mask.

The method is first evaluated through analysis of available commercial masks (Figure 3), including N95 respirators, surgical masks marketed for medical use, and other (in this case, a surgical-style mask with a charcoal-embedded layer marketed for persons with allergies or wearing while exercising in areas with high levels of air pollution). Blue bars show mean particle filtration for masks worn as designed, while gray bars provide a proxy for best possible fit by adding the nylon overlayer. Differences between blue and gray bars provide a measure of the looseness of the fit (extent of leakage of air around the mask in normal wear) while gray bars provide a measure of filtration capacity of the mask material.

**Figure 3:**
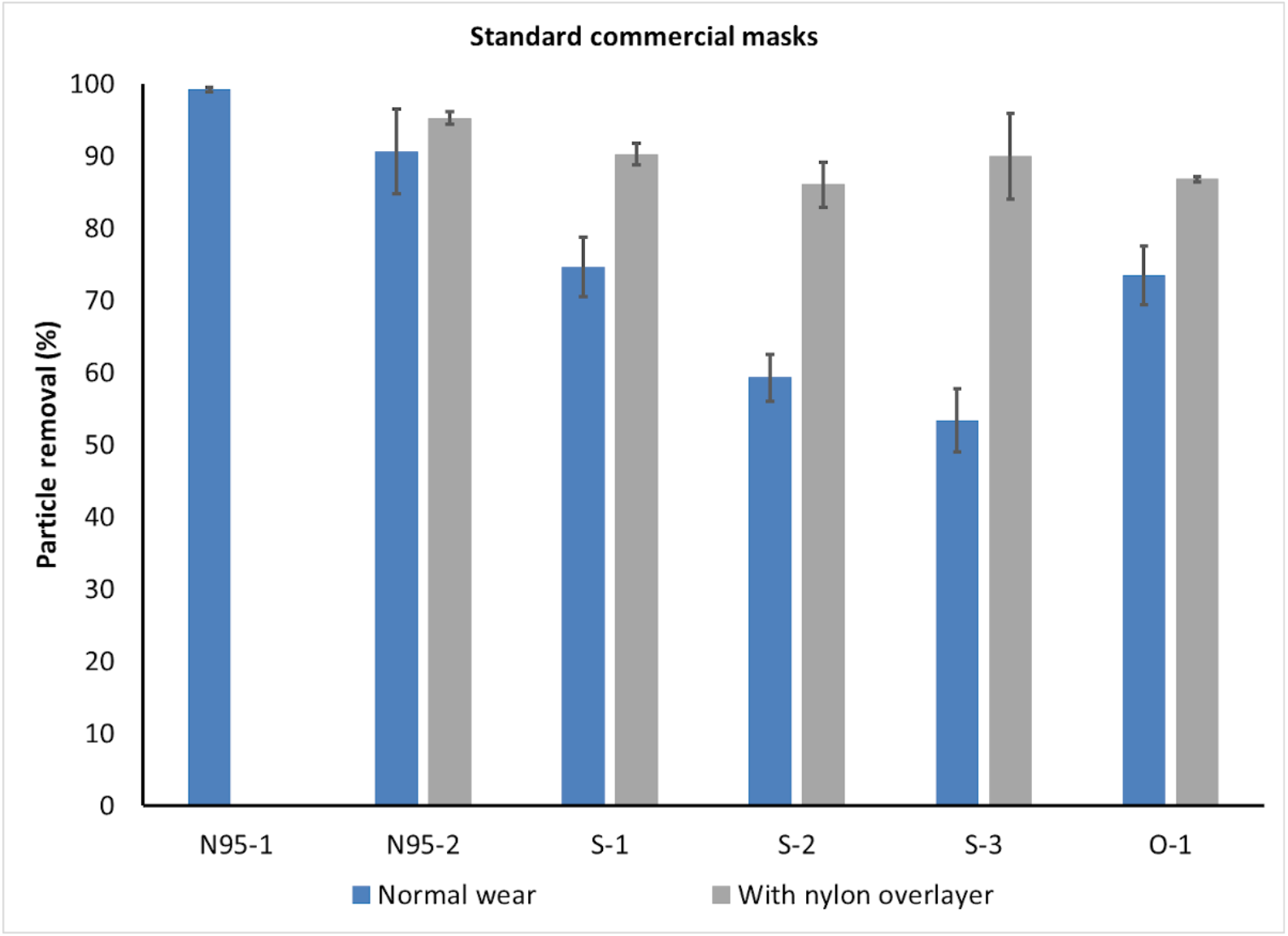
Particle filtration efficiency of standard commercial masks of 3 types: N95 (N95-n), surgical style marketed for medical use (S-n), and Other (O-1, a charcoal filter mask). Data collected with a nylon overlayer holding the mask in place represent a proxy for best-possible fit, i.e., gray bars provide a measure of the filtration capacity of the materials. N95-1 was well-fitted to the mask wearer and shows the expected >99% filtration, while N95-2 was less well fitted, as seen by the difference between the blue and gray bars. While the fit of the three surgical masks (S-1 to S-3) is quite different (blue bars), the materials are comparable (gray bars). Error bars show standard deviation between replicates (n=3 masks for each type tested).

As expected, the mean removal efficiency for the well-fitted N95 mask (N95-1) is greater than 99% with very low variability between replicates (s=0.36%) and low time-based standard deviation (s_t_=0.78%, see Figure 2 data point with highest filtration efficiency). This corresponds to a Fit Factor (C_outside_/C_inside_) of 126, which is above the minimum passable standard of 100^14^, however presentation of results as mean and variability provides more information on the range of particle filtration efficiencies experienced by the user. The poorly-fitted N95 mask (N95-2) has a lower mean removal efficiency 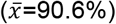, higher variability between replicates (s=5.9%), and higher time-based standard deviation (s_t_=4.6%). This corresponds to a fit factor of 10.6, which is below the minimum passable standard.

In comparison, the standard medical-type masks (S-1 to S-3), when worn over the chin and with an adjusted nose wire, had a mean removal efficiency of only 50 to 75% when worn as designed. In comparison, when tightly fitted to the face using a nylon overlayer these masks achieve from 86 to 90% mean removal efficiency, indicating that (1) the material can actually provide much better filtration than is achieved in normal wear and (2) differences between brands are primarily in the quality of fit rather than the quality of material used. Interestingly, in this case the carbon filter mask (O-1) performs approximately as well as the best performing surgical mask despite a significant difference in the design specifications and materials used.

The same measurements and metrics were then used to test fifteen different cloth masks being made or marketed to the public at this time (April-May 2020). Results (Figure 4) are presented as absolute particle removal efficiency (top panel) and in comparison with the top performing surgical mask (S-1) (bottom panel). While these masks represent a small subset of available masks and materials, several useful preliminary observations can be made.

**Figure 4:**
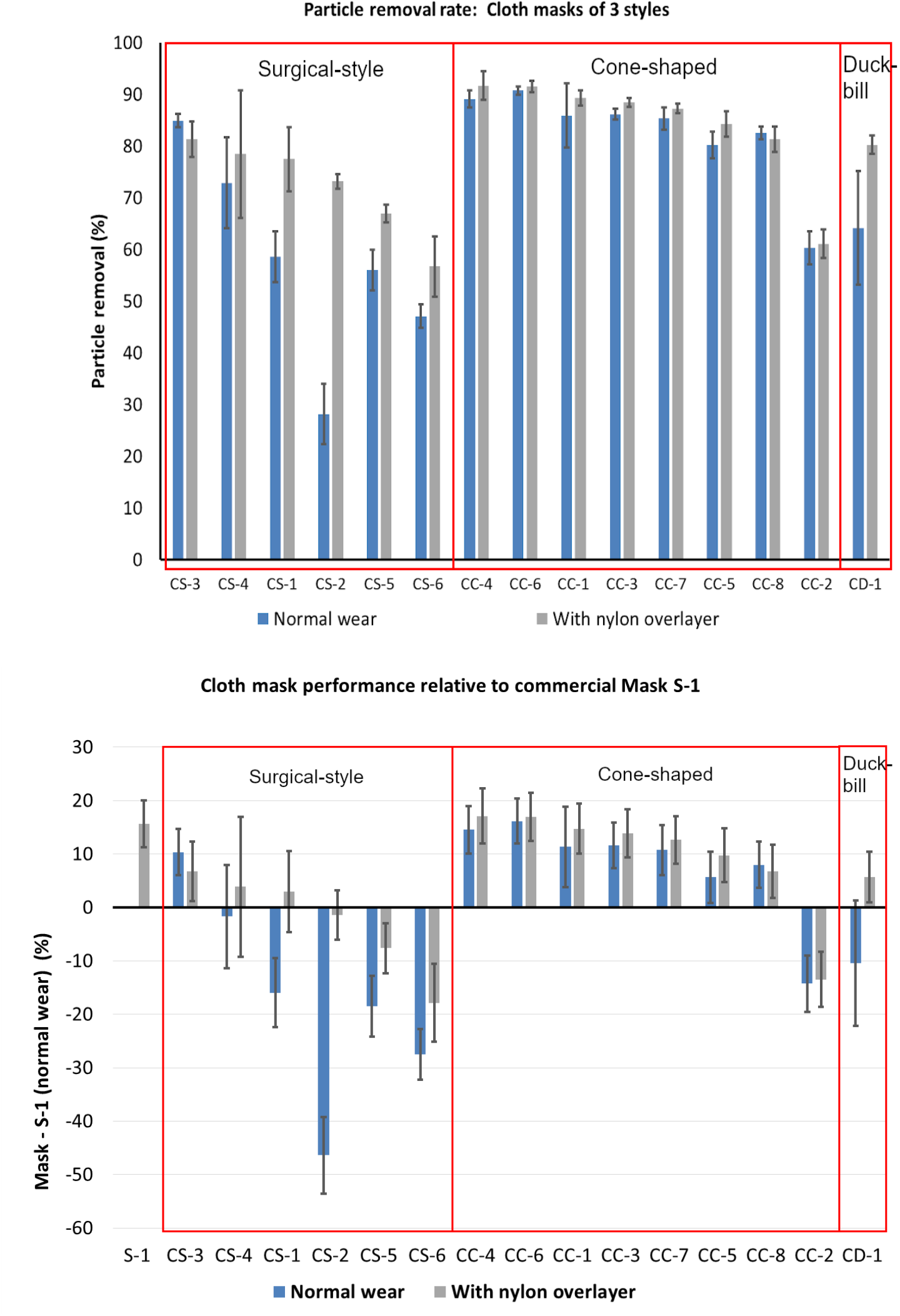
Performance of a range of cloth masks being made by the community and by commercial vendors presented as absolute performance (top panel) and in comparison to S-1, the top performing surgical mask (bottom panel). Preliminary data show the difference between performance of masks using different form factors, e.g., cone-shaped masks appear to have a better and more consistent fit to the face. Notably multiple cloth masks perform as well as or better than surgical masks when worn as designed, and some provide equivalent filtration to surgical masks snugged to the face. However, there is wide variability in filtration provided by cloth masks, due both to fit (difference between blue and gray bars) and materials (gray bars).

First, quality of cloth masks is highly variable, both in fit (difference between blue and gray bars) and material filtration capacity (gray bars); therefore the public would greatly benefit from a quantitative method for evaluating masks they may be considering for health protective reasons. Second, it appears that different masks shapes may provide a more consistent fit even when hand-made using standard patterns; e.g., in these data the cone masks appear generally to fit better than the surgical-style masks (as evaluated by difference between blue and gray bars, where addition of the nylon layer generally improved performance for surgical-style masks but not for cone-shaped masks). Exceptions to improvement when adding the nylon overlayer were rare and due to material stiffness where the mask could not completely conform to the wearer’s face and therefore the nylon layer led to bunching (creation of new air leakage pathways). The nylon layer also reduced the variability with time as indicated by a decrease in the time-based standard deviation. Both of these metrics indicate improved protection for the wearer from particle inhalation.

When using mask S-1 worn as designed as a baseline, several of the cloth masks match or exceed this performance (Figure *4*, bottom panel). The masks that achieved this level of filtration without the stocking overlayer were cone shaped and included a layer of meltblown filter fabric, similar to interfacing layers being added to many homemade masks, and specified as BFE85, between fabric cover layers. Additional filter layers including water-repellent non-woven cloth marketed as disposable massage table covering and dry disposable baby wipes, improved the filtration efficiency only moderately in the cone shaped masks. Surgical-style masks that achieved the best filtration efficiency with the addition of a nylon overlayer included a filter layer (organic cotton batting, Pellon, or loosely-woven cotton muslin) between two layers of cotton fabric. These data are not included here due to limited number of replicates, but are available on a web portal (SI).

## 3. Conclusion

A rapid testing protocol is presented for evaluation of loose-fitting type masks to provide quantitative, intercomparable data for particle removal efficacy of masks made with different types of fabrics and with different designs/fits, independently providing an assessment of the quality of the mask fit and the material used. The protocol collects high-resolution particle count data inside and immediately outside of masks to report both mean and time-based standard deviation of particle removal efficiency, while wearing the mask as-designed and under a nylon layer that snugs the mask to the face. The protocol is validated on a well-fitted N95 mask, and a commercial surgical-type mask is used as a reference baseline for evaluation of alternative mask particle removal efficiencies. Commercial surgical masks marketed for medical use had mean particle removal efficiencies from 50-75% when worn as designed but up to 90% when snugged to the face under a nylon layer. Cloth masks tested had widely varying mean particle removal efficiencies (<30% to near 90%), with some cloth masks achieving similar filtration efficiencies as commercial surgical masks. However, in general, surgical-style cloth masks had poor fit (i.e., performance was greatly enhanced with the nylon overlayer) compared to cone-shaped masks, and masks with good material filtration performance tended to have a filter layer (e.g., meltblown BFE85 filter layer) in addition to two layers of cotton or non-woven fabric.

This rapid testing method (~30 minutes per mask design including replicates for statistical validity) provides a holistic evaluation of mask particle removal efficacy (material, design, and fit) while enabling independent evaluation of these characteristics. This method uses instrumentation that is typically available in many health centers and fire stations, as well as other facilities, and compensates for assumptions made in fit-testing programs so that it can be easily replicated for on-site testing of specific masks across many communities.

## 4. Experimental Procedures

### Particle counters

Particles in ambient air and air inside of the mask breathing zone were counted using two PortaCount Plus Model 8028 instruments running in count mode. The PortaCount Plus instrument uses a condensation particle counter to determine particles per cm^3^ in air sampled at a flow rate of 1.67 cm^3^/s and reports one value (in particles/cm^3^) each second.^15^ The instrument counts particles ranging in size from 0.02 to >1 μm, however data on the size distribution of counted particulates is not reported; size distribution of the particles used to challenge the masks was therefore measured independently, as reported in the following section.

Mask fit testing is usually conducted using a single instrument in fit test mode, which sequentially tests air inside and outside the mask and therefore depends on an assumption of consistent particle concentration in the room. As this assumption was frequently violated in our test setup even in cases where particle generation was used (which may therefore commonly be the case in other facilities), here two PortaCount instruments were used in count mode to simultaneously collect and display continuous data for ambient air and inside mask air at high frequency (1 Hz) during minute-long tests.

**Figure 5:**
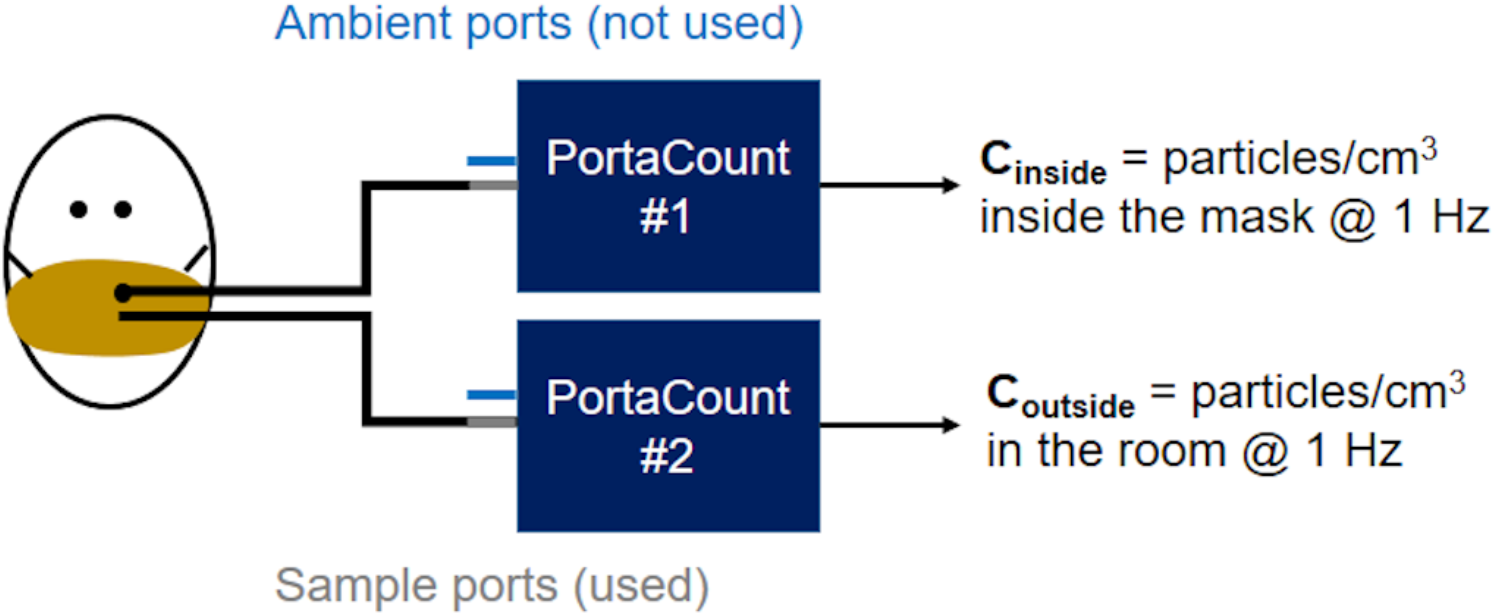
Test setup has two PortaCount systems running in parallel; 1/8” diameter tubing of identical length connects (top) the mask port to one instrument sampling port and (bottom) a tube inlet located just outside the mask to the second instrument sampling port. Both instruments are run in “Count” mode where concentrations are reported once per second (1 Hz). The dual-instrument configuration is required because each instrument has only one internal measurement cell, for which the input is swapped between the sampling and ambient inputs during standard fit testing. Photograph of PortaCounts used, including ports and tubing available in SI (Figure S1).

Two 1/8” ID tubes (sold with the PortaCount Instrument) trimmed to equal length (approximately 100 cm) sampled air just inside and outside of the mask. Air inside the mask was sampled through a tight-fitting grommet inserted into each mask using a TSI Fit Test Probe Kit (model 8025-N95) and positioned at the philtrum of the upper lip per standard mask testing guidance appropriate to the shape of each mask. Ambient air was sampled from a position ~3 cm from the grommet on the outside of the mask.

### Particle Generation and Characterization

All tests were run in a 65 m^3^ rectangular room after at least 15 minutes of operating a TSI Particle Generator Model 8026 (TSI Incorporated, Shoreview, MN, USA). This tool is typically used in conjunction with TSI PortaCount instruments to ensure sufficiently high particle counts and appropriate size distributions to meet OSHA standards. Particles were generated from a dilute (2%) solution of sodium chloride (NaCl), reported to have a nominal size of 40 nm with a geometric standard deviation of 2.2 based on instrument specifications.^16^ To verify that the particles challenging the masks was comprised primarily of these generated particles, the particle size distribution in the room was characterized by running three 5-minute tests approximately hourly on several testing days (n=21 in replicates of 3) using the TSI Engine Exhaust Particle Sizer Spectrometer (EEPS) Model 3090 (for particles in the range 5.6 to 560 nm, with 32 channels acquired at 10 Hz) and the TSI Optical Particle Size Spectrometer (OPS) Model 3330 (for particles in the range 0.3 to 10 μm, with 16 channels acquired at 1 Hz). The size distribution of particle number concentration was consistent at all times and days sampled, which supported the averaging of collected data. The histogram of average normalized particle frequency revealed a bimodal distribution of particle sizes, shown in Figure 6. Confidence intervals (CI) for the count median diameter (CMD) and the geometric standard deviation (GSD) were calculated from Student’s t-test statistics, with p=0.05 and M=20 degrees of freedom. The first peak likely represents particles that are not filtered by building HVAC systems, as the distribution parameters (CMD=9.53 ± 2.19 nm, GSD=1.23 ± 0.13; average ± 95% CI) are consistent with background air measurements reported by the authors in other rooms and buildings on campus. Features of the second peak (CMD = 37.30 ± 15.40 nm, GSD = 1.79 ± 0.44) are in agreement with specifications reported for the TSI Particle Generator manual. Overall, 97.01 ± 0.02% (average ± 1·s) of particles are in the standard range used to challenge masks (<300 nm), so the reported particle filtration efficiencies can be directly compared to numbers reported to comply with OSHA standards.

**Figure 6:**
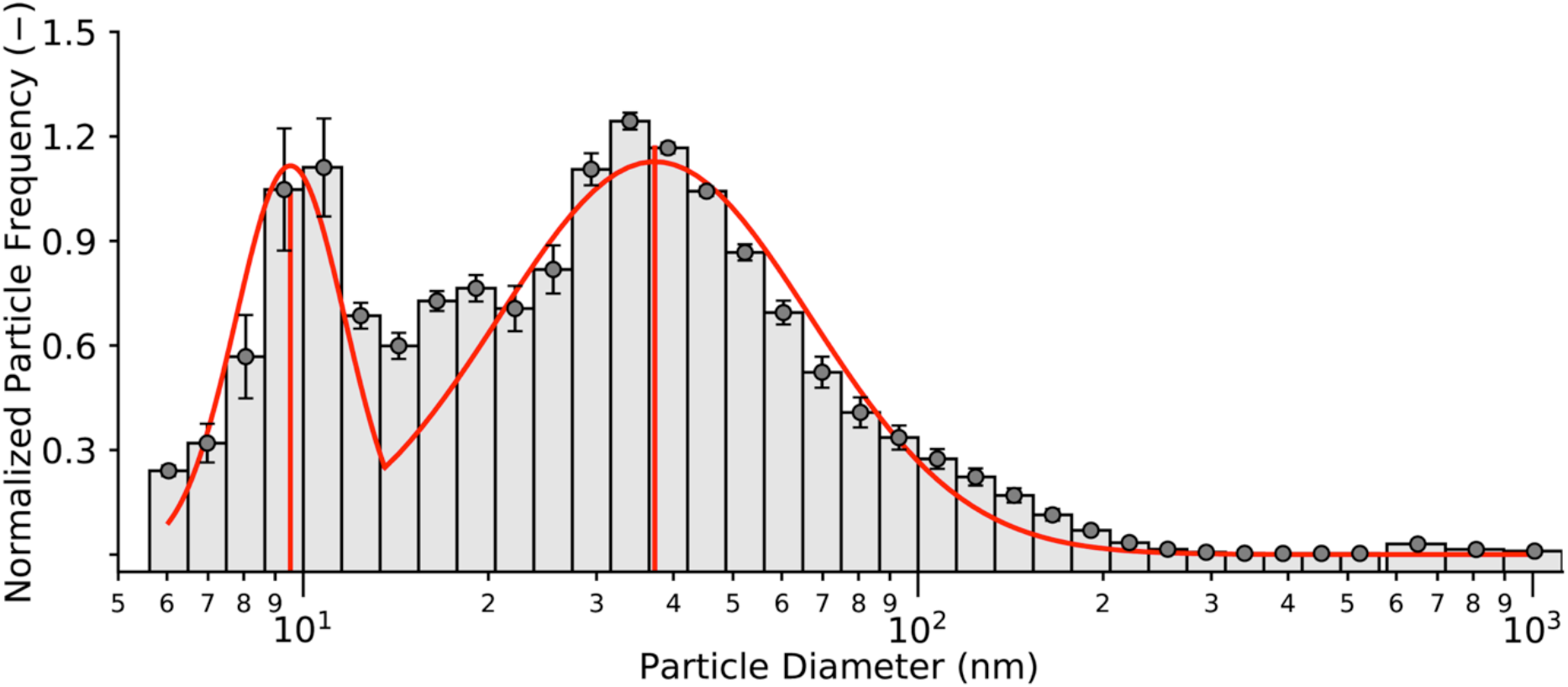
Average histograms of normalized particle frequency as a function of size, with superimposed bimodal lognormal distribution. Count median diameter (CMD ± 95% CI) is 9.53 ± 2.19nm for the first peak and 37.30 ± 15.40nm for the second peak. Geometric standard deviation (GSD ± 95% CI) is 1.23 ± 0.13 for the first peak and 1.79 ± 0.44 for the second peak. Particles generated by the TSI Particle Generator account for the larger peak, while particles in lab air account for the smaller peak.

### Calibration

An inter-calibration was conducted between the two PortaCount modules to account for any drift or changes in calibrations due to, e.g., wick saturation. Each sampling day, calibration data (a minimum of three one-minute time series, n=180) were collected by recording readings simultaneously on both instruments while sample tubes were side-by-side (within 3 cm), open to the air (no mask), and a minimum of 1m from any person and 2m from the particle generator (as recommended by the manufacturer). Correlation coefficients between the readings from the two instruments were consistently above 0.9, and day-specific linear regressions were used to normalize particle counts from the Reference PortaCount to equivalent particle counts from the Mask PortaCount before calculating particle removal efficiencies.

### Data collection and processing

Each mask test consisted of three one-minute runs while wearing the mask as designed (Figure *7* (a)). In addition, the mask material was held against the face by adding a section of nylon stocking over the entire mask area following recommendations from Copper et al.^9^ (Figure *7* (b)) to simulate best possible fit and provide information on material filtration, and a single one-minute test was recorded in this configuration. All masks except the well-fitted N95 were tested in this second configuration. Results are reported only for masks for which at least three replicate sample masks were available (data for masks with n<3 are being provided through our web portal).

Particle concentration data from inside and outside the mask was logged each second for the one minute tests using video capture and subsequently transcribed to a database (noting that newer PortaCount models can log count data through a software interface to simplify data collection). Particle removal at each time step was calculated as follows:

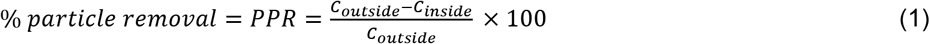

where *C_outside_* is the corrected reading from the Reference PortaCount (as described above) and *C_inside_* is the reading in the breathing zone of the mask.

Average particle removal efficiency (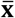, reported as % removal), standard deviation between masks (generally n=3, ***s*** reported as %), and mean standard deviation over the one-minute tests (***s_t_***, reported as %) were computed for each mask with and without a nylon stocking layer. These summary statistics can be used to calculate Fit Factor for the masks, if desired, using Eqn. 2:

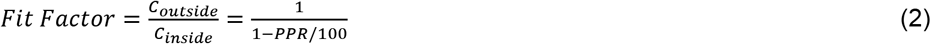

**Figure 7:**
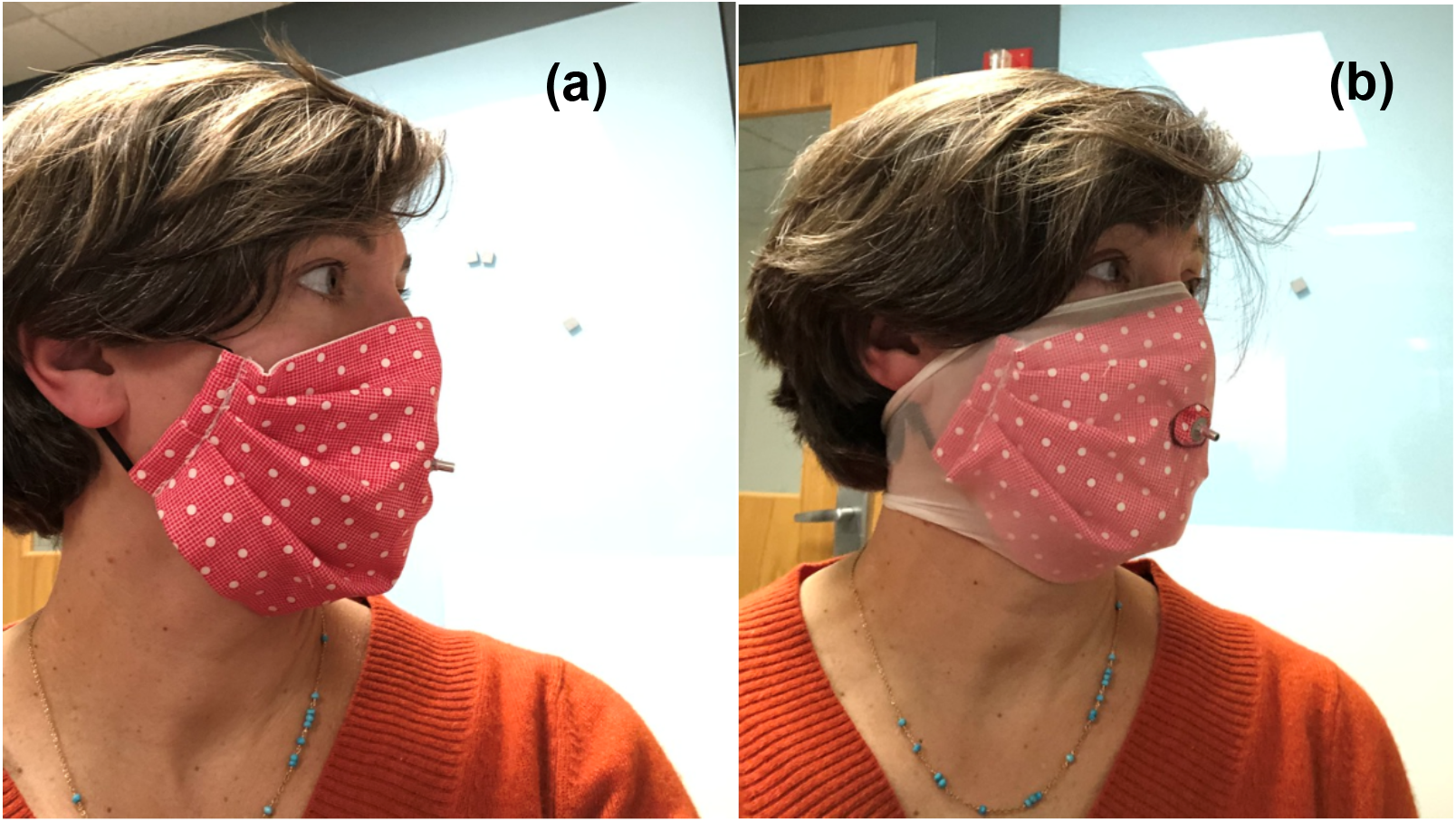
Facemask (Mask CS-1) worn as designed **(a)** and with a nylon stocking layer **(b)** with tightly-sealed grommet positioned at the philtrum of the upper lip. The grommet is used to sample air from inside the mask during testing. Note: this mask could have been worn inside-out to ensure the folds faced down. However, for the purposes of this test precautions against particle collection in folds were not considered necessary.

### Masks

Masks tested are given labels according to the mask type and then an individualized sample number. Commercial masks are divided into N95-type (N95-1, N95-2, etc.), surgical-style (S-1, etc.), and other (O-1, etc.). Cloth masks are given a pre-pended “C” identifier and divided into surgical-style (CS-1, etc.), cone-shaped (CC-1, etc.), and duck-bill shaped (CD-1, etc.) (Figure S2). Results are reported for a range of commercially-produced, medical-type facemasks (masks with elastic ear loops and in-sewn wires to adjust fit to the bridge of the nose), and fifteen sewn fabric facemasks of various designs that were sourced from community volunteers producing masks for essential personnel as well as online vendors that have started to market masks of this type since March 2020 (Table S1). Several of the fabric masks included filter layers such as non-woven polypropylene fabric, meltblown textiles, and disposable baby wipes. In addition, several sewn masks included hydrophobic layers including interfacing (Pellon) and non-woven fabric marketed as disposable massage table covering. Some masks included wires to fit the masks across the bridge of the nose. A set of well-fitted (N95-1) and poorly fitted (N95-2) masks were tested to validate the protocol.

## Data Availability

Data referred to in this manuscript is available upon request from the authors.

## Acknowledgements

Funding for this work was provided by Northeastern University Office of the Provost through a COVID-19 Seed Grant. Advice, materials, tools, instruments, and time were donated or shared with us in order to carry out this project. A critical TSI PortaCount model 8028 was donated by Pfizer Inc. through the coordination of Luanne Kirwin, Sonya Ross, Rob Silk, John Price, and Michael Glover. Tools, advice, and expertise in conducting fit testing were generously provided by Gregory Lawless, TSI Incorporated; Albert Thomas, the Massachusetts Department of Fire Services; George Mulholland, Department of Chemical and Biomolecular Engineering, University of Maryland; Marco Fernandez, National Institute of Standards and Technology; and John Price and Dan Meinsen, Environmental Health and Safety, Northeastern University. Jessica Benedetto, Laurie Bowater, Jennifer Cawley, Benjamin Chung, Steve Gransbury, Denise Hajjar, Michelle Kane, Laura O’Neill, Waas Porter, Stephanie Potts, John Price, Alan Raymond, Renée Scott, and Mark Vera kindly donated masks. Data extraction was provided by dedicated undergraduate students Matthew Biega, Alina Dees, Ariana Patterson, and Kelsey Walak. Thanks also to Christine Trowbridge for assistance in editing.

## SUPPLEMENTAL INFORMATION

**Table SI1.**
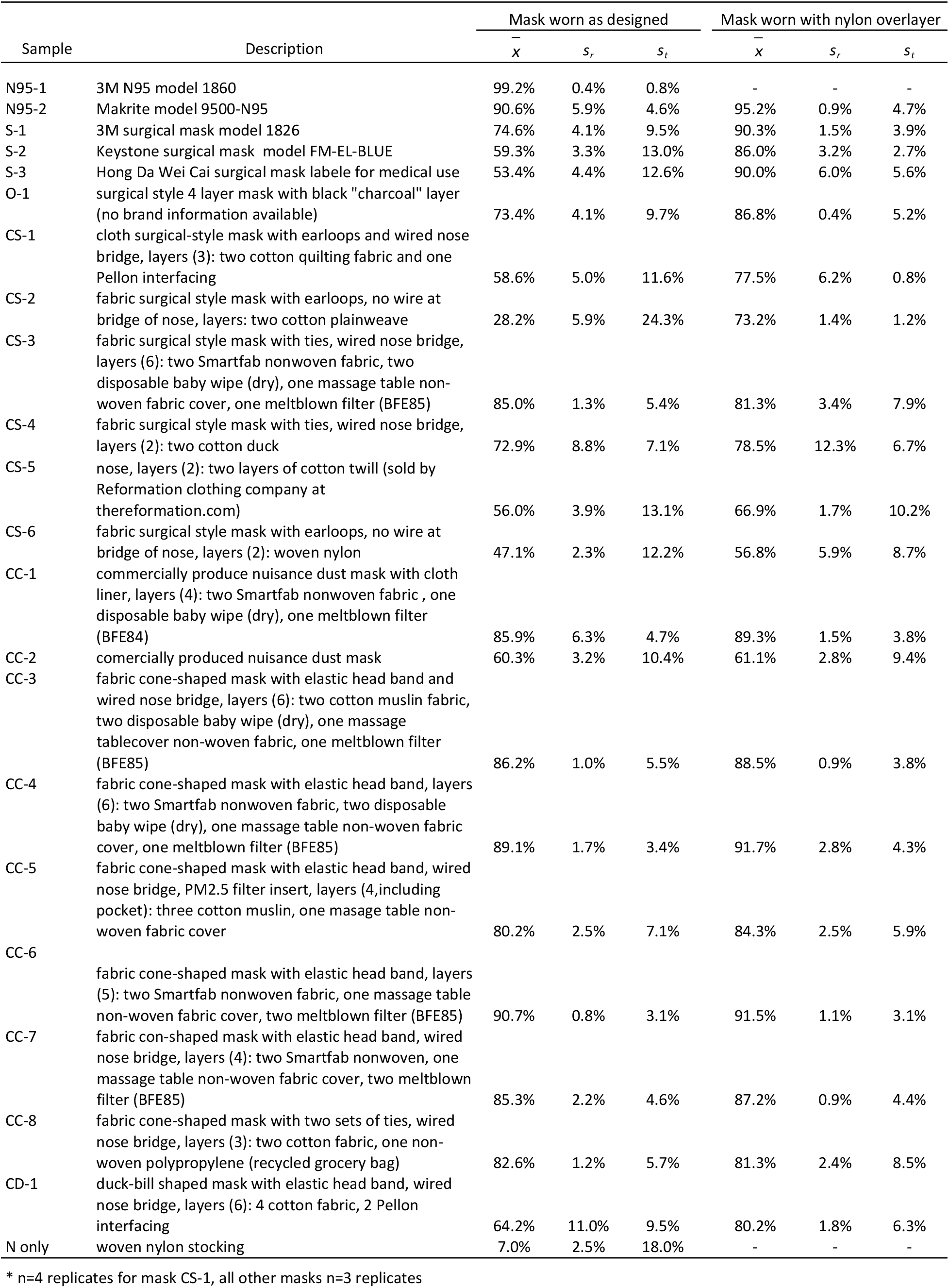
Mask details, mean filtration efficiency 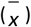, standard deviation of mean filtration efficiency between replicates* (s_r_), and standard deviation of filtration efficiencency over one minute runs (s_t_).

**Figure S1.**
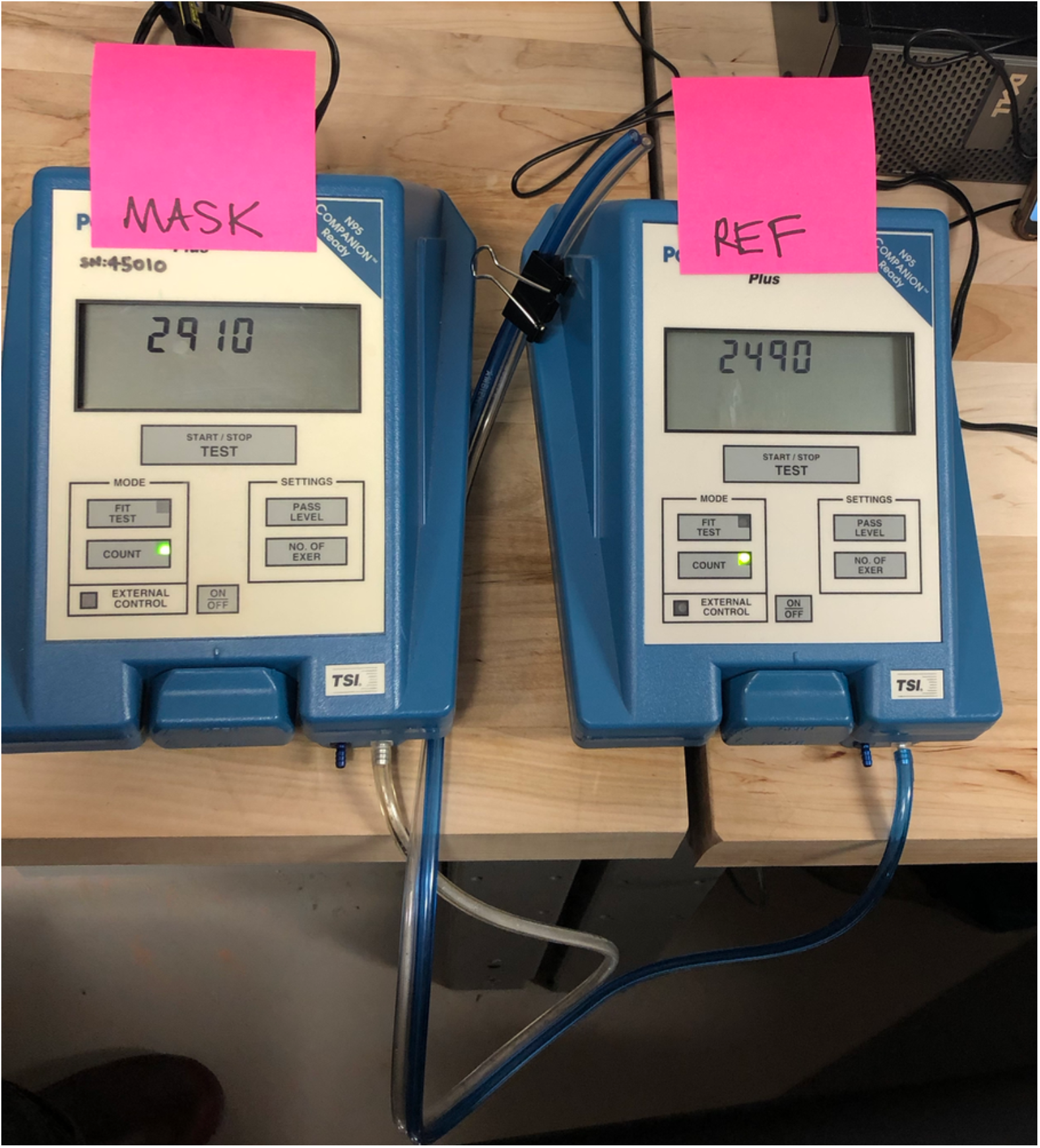
Two TSI PortaCount model 8028 used in this work. Sample tubes are of equal length and are connected to right-hand ports labeled “sample”. Instruments were operated in count mode with “Mask”-labeled instrument sampling air from inside the mask and “Ref”-labeled instrument sampling ambient air just outside of the mask.

**Figure S2.**
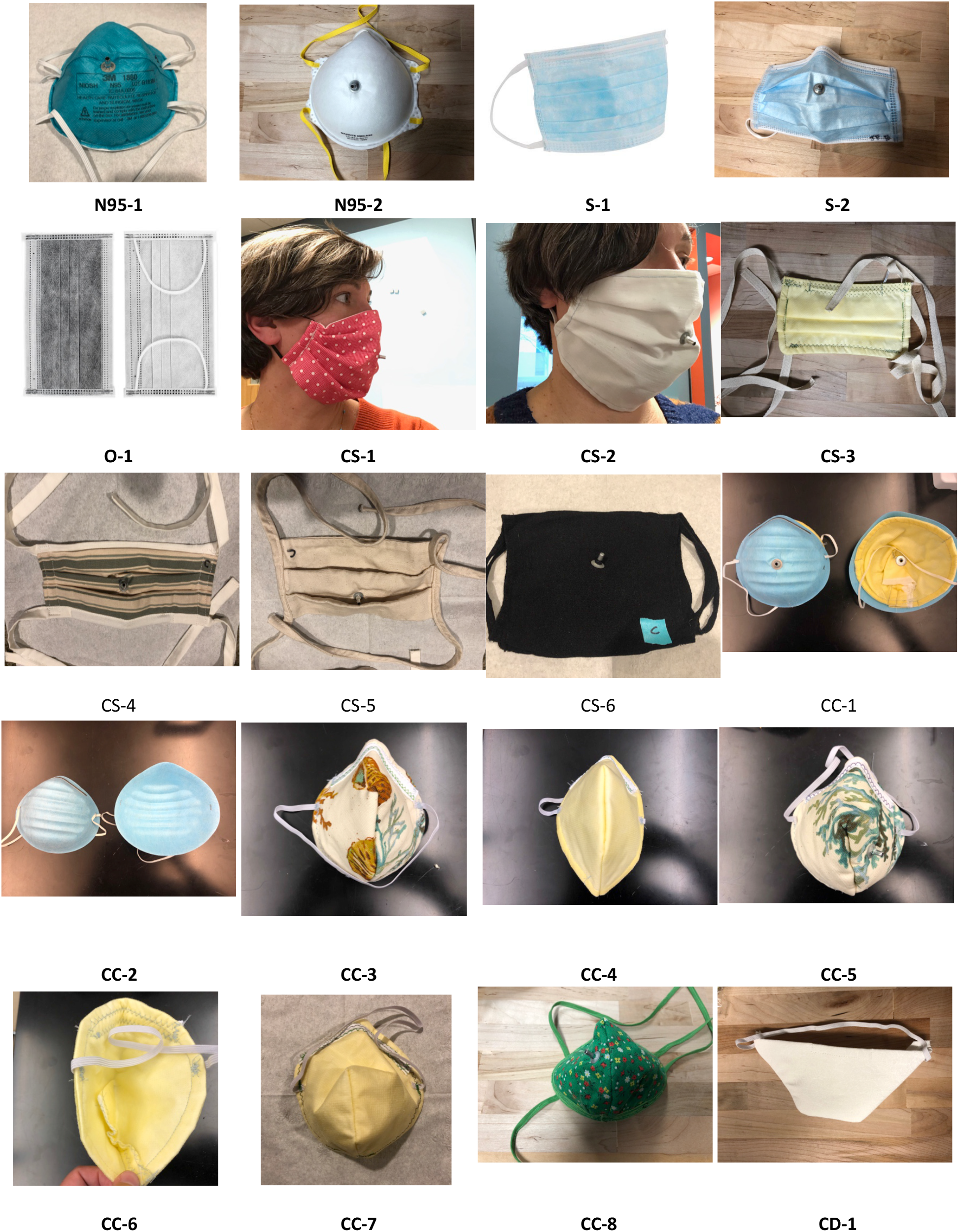
Gallery of mask images. Masks ordered by sample ID. Descriptions included in Table S1. Additional and updated results are available through a web portal at masktestingatNU.com.

